# Polymorphism in IFNAR contributes to glucocorticoid response and outcome in ARDS and COVID-19

**DOI:** 10.1101/2022.03.10.22272123

**Authors:** Juho Jalkanen, Sofia Khan, Kati Elima, Teppo Huttunen, Ning Wang, Maija Hollmén, Laura L. Elo, Sirpa Jalkanen

## Abstract

The use of glucocorticoids has given contradictory results for treating acute respiratory distress syndrome (ARDS). Here we report a novel disease association of a SNP rs9984273, which is situated in the interferon alpha/beta receptor (IFNAR2) gene in an area corresponding to a binding motif of the glucocorticoid receptor (GR). The minor allele of SNP rs9984273 associates with higher IFNAR expression, lower IFN-gamma and IL-6 levels and less severe form of coronavirus diseases (COVID-19) according to the COVID-19 Host Genetics Initiative database, and better outcome in interferon (IFN) beta treated patients with ARDS. Thus, the distribution of this SNP within clinical study arms may explain the contradictory results of multiple ARDS studies and outcomes in COVID-19 concerning type I IFN signalling and glucocorticoids.

**One-Sentence Summary:** Single nucleotide polymorphism in interferon receptor contributes to corticosteroid response and outcome in ARDS and COVID-19

---

**I**nterferon (IFN) and cortisol are our natural responses to viral insult and stress, respectively, and the inability to summon either is deadly *(1-3)*. It appears that these natural responses need to happen in a sequential manner, since endogenous cortisol and given glucocorticoids block IFN signalling *(4-6)* and are known to be harmful if given early in viral diseases *(7)*. For decades in the clinical management of ARDS, sepsis, and now COVID-19, glucocorticoids have given controversial results and their use still remains an ongoing debate *(7-13)*. The current SARS-CoV2 pandemic has again showed us, that the early use of glucocorticoids in a severe viral respiratory infection is harmful, but late in the disease when excessive inflammation and poor oxygenation prevails glucocorticoids are beneficial *(14)*. In addition to glucocorticoids, Type I IFNs are also considered as viable treatments for COVID-19, Middle East respiratory syndrome (MERS), and ARDS, but contrary to glucocorticoids IFNs should be administered early *(15-17)*. Given late in the disease course of COVID-19 or in non-COVID-19 ARDS, the combination of intravenous IFN beta and glucocorticoids has even shown to be harmful *(5, 18, 19)*. Hence, we hypothesize that nature has never intended these two fundamental pathways to be “on” at the same time. In our work we have come across the SNP rs9984273, which is situated in the gene of IFNAR2. Its location corresponds to a binding motif for GR. It appears that this SNP, at least to a certain extent, blocks glucocorticoid interference to type I IFN signalling, thereby enabling us to take advantage of these two pathways. The SNP is associated with a milder disease in COVID-19 and better survival in patients with ARDS when glucocorticoids are used together with IFN beta. Here we elucidate our findings with the help of clinical studies and the publicly available COVID-19 GWAS Task Force data.

## Discovery of rs9984273 in clinical trials investigating intravenous IFN beta-1a for the treatment of ARDS

We have performed two significant clinical trials investigating the use of intravenous IFN beta-1a for the treatment of ARDS *(18, 20)*. The rationale for performing these trials was in the ability of intravenous IFN beta to increase the expression of 5’-ectonucleotidase, CD73 on lung endothelium. CD73 is a key protective molecule and the rate limiting enzyme in converting pro-inflammatory adenosine monophosphate (AMP) into anti-inflammatory adenosine *(21)*. The first open-label Phase I/II study investigating intravenous IFN beta-1a for ARDS produced remarkable results, showing ARDS mortality of only 8% *(20)*. The trial was small, consisting of only 37 ARDS patients in the active arm, but it led us to perform a substantially larger placebo controlled trial of 296 ARDS patients *(18)*. In this later randomized clinical trial intravenous IFN beta showed no benefit over placebo in the entire study population. However, further laboratory analyses revealed that glucocorticoids block IFN signaling and the up-regulation of CD73 in the lung endothelium and increased mortality in the active arm of the study *(5)*. Patients (n=66) who did not receive glucocorticoids with IFN beta had 28-day mortality of 10,6%, while patients (n=78) who did receive glucocorticoids with IFN beta had 28-day mortality of 39,7%. The deleterious effect of glucocorticoids given together with intravenous IFN beta was evident despite adjusting to disease severity and the likelihood of receiving glucocorticoid treatment in propensity matched multivariable regression analyses *(5)*.

Further, pre-defined targeted resequencing and genetic analyses of entire gene regions were performed for genes encoding CD73 *(NT5E)*, IFN alpha/beta receptor alpha and beta chains *(IFNAR1* and *IFNAR2)*, Minichromosome Maintenance Complex Component 2 *(MCM2)*, Angiopoietin 2 *(ANGPT2)*, and Ras Association Domain Family Member 3 *(RASSF3)* genes. Out of these analyses rs9984273 was shown to significantly associate with mortality in the IFN beta treatment arm after adjusting for multiple testing. Patients with ARDS carrying the minor C allele had day-28 mortality of only 10,6% despite receiving glucocorticoids with IFN beta. A similar phenomenon was not witnessed in the placebo arm (Fig. 1A and B). Multivariate regression modelling adjusting for disease severity according to the pre-dose Acute Physiology and Chronic Health Evaluation (APACHE) II score showed that concomitant use of glucocorticoids with IFN beta-1a was associated with increased mortality (OR 3.30; 95% CI, 1.79 – 6.08; P < 0.001), while the presence of the minor allele C in rs9984273 was associated with lower mortality at day 28 (OR 0.31; 95% CI, 0.13 – 0.72; P = 0.006) (Fig. 1C). Thus, the use of glucocorticoids and the presence of the minor allele in this SNP dictate the outcome of intravenous IFN beta treatment in patients with ARDS. This finding was further validated by retrospectively performing focused genetic analyses of IFNAR2 on the Phase I/II study population. Phase I/II and III study populations were comparable according to demographics and disease severity; however, we identified one major difference. In the Phase I/II study 40% of the study subjects receiving IFN beta received overlapping glucocorticoids, but 73% of these patients had the rs9984273 C allele. In contrast, in the phase III study up to 60% of patients received corticosteroids, but only 40% of the subjects receiving overlapping glucocorticoids with IFN beta-1a carried the minor allele. Hence, the rs9984273 C allele was highly enriched in the Phase I/II study population compared to the Phase III study, and more patients received glucocorticoids in the Phase III than in the Phase I/II study.

**FIG. 1.**
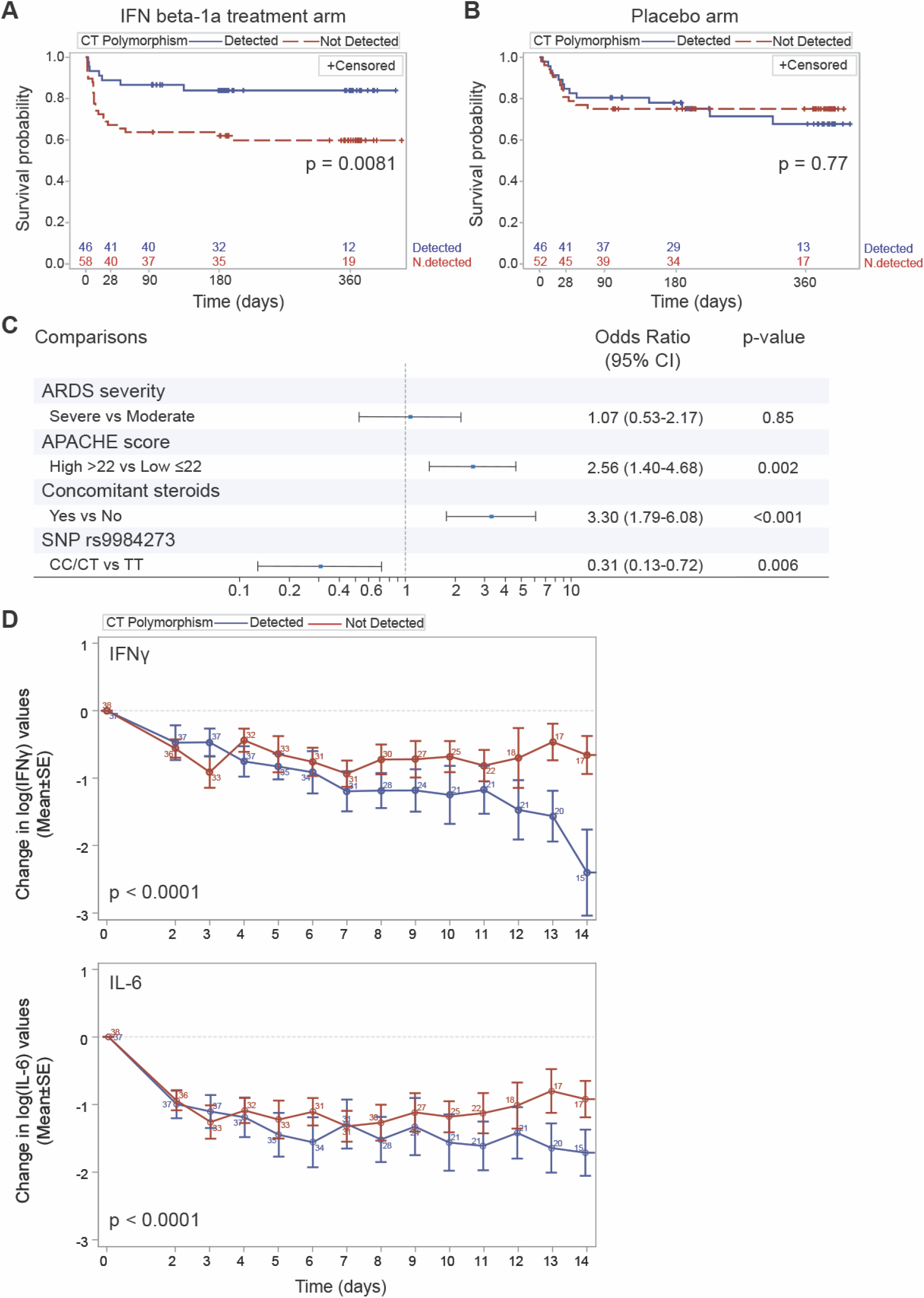
The effect of the rs9984273 genotype on mortality and cytokine levels in the INTEREST trial investigating intravenous IFN beta-1a for ARDS treatment. A genetic sample was available from 202/296 (104/144 in the IFN beta-1a treatment arm and 98/152 in the placebo arm) of the study subjects. Number of the subjects at risk is indicated on the x-axis. (**A)** The rs9984273 minor allele C (CC or CT) was significantly associated with survival in patients compared to those who were homozygous with the major allele (TT). (**B)** A similar association was not observed in the placebo arm. (**C)** Further non-adjusted sub-group analyses encompassing the entire Phase III study population (both active and placebo arms) revealed that the use of glucocorticoids had the strongest association with altering the treatment effect of INF beta-1a. **(D)** The logarithmic change in serum IFN gamma and IL-6 levels according to the genetic background of rs9984273 in patients of the INTEREST trial with ARDS of pulmonary origin. Patients homozygous with the major allele (TT) compared to the minor allele (CT or CC) of rs9984273 show higher serum IFN-gamma and IL-6 levels when given glucocorticoids.

## Rs9984273 associates with IFN gamma and IL-6 production in patients receiving glucocorticoid treatment

The SNP rs9984273 locates in the 3’ untranslated region (3’-UTR) of the IFNAR2 gene (chr21:33262760 (GRCh38)). To investigate potential binding sites of GR in the region, we utilized position weight matrices compiled from individual genomic sites from the Transfac database (Transfac matrix table, Release 2020.3) or derived from the Encode ChIP-seq data for GR *(22)*. The Transfac based results showed that the SNP resides in a predicted transcription factor binding site of GR (Transfac matrix similarity score 0.864, chr21:33262746 – 33262761). Furthermore, the SNP was predicted to alter a GR binding site on the basis of the ChIP-seq data (GR_known3 from *(22)*).

Following our *in silico* evaluations, we next wanted to determine, whether rs9984273 was associated with the immune status of the patients when given glucocorticoids. The Phase III ARDS trial (INTEREST) was optimal to analyse this effect as it had cytokine profiling of the patients for the first 14 days. IFN-gamma and IL-6 were chosen as indicators of immune activation as they are also associated with poor outcome in ARDS and COVID-19. Only those patients with ARDS of pulmonary origin (pneumonia or pulmonary sepsis) were chosen for these analyses to best resemble ARDS of COVID origin and adjust for a heterogenic etiology of an all-comer ARDS trial. We found that patients homozygous with the major T allele of rs9984273 have approximately the same level of IFN gamma as those with the minor allele C (CC or CT) until day 7, but thereafter patients with the TT genotype started to show higher IFN-gamma and IL-6 levels compared to the patients with the CC/CT genotype (Fig. 1D). Importantly, this finding indicates that administering patients with glucocorticoids is not harmful, if they possess at least one copy of the minor allele despite receiving simultaneous IFN beta; however, for patients homozygous with the major allele TT, glucocorticoid use is associated with higher levels of IFN gamma and IL-6 (RMANCOVA, P < 0.0001), and the two anti-inflammatory agents glucocorticoids and IFN beta become inflammatory.

## Rs9984273 contributes to the expression of IFNAR

After the discovery of rs9984273, and recognizing its role in the outcome of the previous clinical ARDS studies, we then investigated, whether this polymorphism contributes to the expression of the IFNAR (composed of IFNAR1 and IFNAR2 subunits) by immunohistochemistry using an antibody against IFNAR1 subunit of the receptor in surgical lung specimens from 14 patients. In this material 5 samples had the major allele TT, 8 samples CT and 1 CC for rs9984273. The primary antibody was titrated to the level, at which the alveolar epithelium and vasculature still remained strongly positive in a subset of the samples. The scoring was done without any knowledge of the genotype. The results clearly show that all high expressors of IFNAR were CT:s and overall the CT/CC group had statistically significantly higher IFNAR but four of nine samples from the CC/CT group show the same expression level as the TT group, indicating that other factors are involved in regulating IFNAR expression.

## CT and TT patients display different features in IFN beta-induced STAT1 and STAT2 signaling and responsiveness to glucocorticoids

As the immune status and IFNAR expression are at least partially under the control of rs9984273 and the formation of the signal transducer and activator of transcription 1/2 and interferon regulatory factor 9 (STAT1/STAT2/IRF9) complex and its translocation into the nucleus is required to trigger type I IFN responsive genes, we next examined STAT1, phosphorylated STAT1 (pSTAT1) and STAT2 expression in the lung specimens cultured in the presence of IFN beta with or without hydrocortisone (HC). Glucocorticoids have been reported to inhibit this complex formation and its nuclear localization, and thus IFN beta signaling *(4, 5)*. We found that TT patients tended to have lower total STAT1 expression and statistically significantly less pSTAT1 than the CT patients and HC inhibited its nuclear location (Fig. 3, A and B). In contrast, TT patients had significantly more STAT2 than the CT patients and HC did not decrease its nuclear expression like in CT patients (Fig. 3, C and D). It has been shown that STAT2 upregulates IL-6 via co-operation of the NF-KB pathway *(23)*. In this scenario, we speculate that the ‘extra’ STAT2 in patients with the rs9984273 TT genotype maintains the high IL-6 and IFN-gamma levels presented in Figure 1D via a comparable mechanism. For the STAT-analyses, we were limited being able to only include three out of five CT patients to the combined data shown in Figure 3, as two sample donors were on corticosteroids at the time of the operation. In samples of those two donors, IFN beta could not induce efficient nuclear localization of STAT1 and STAT2, further demonstrating the clinical role of glucocorticoids in this phenomenon.

**FIG. 2.**
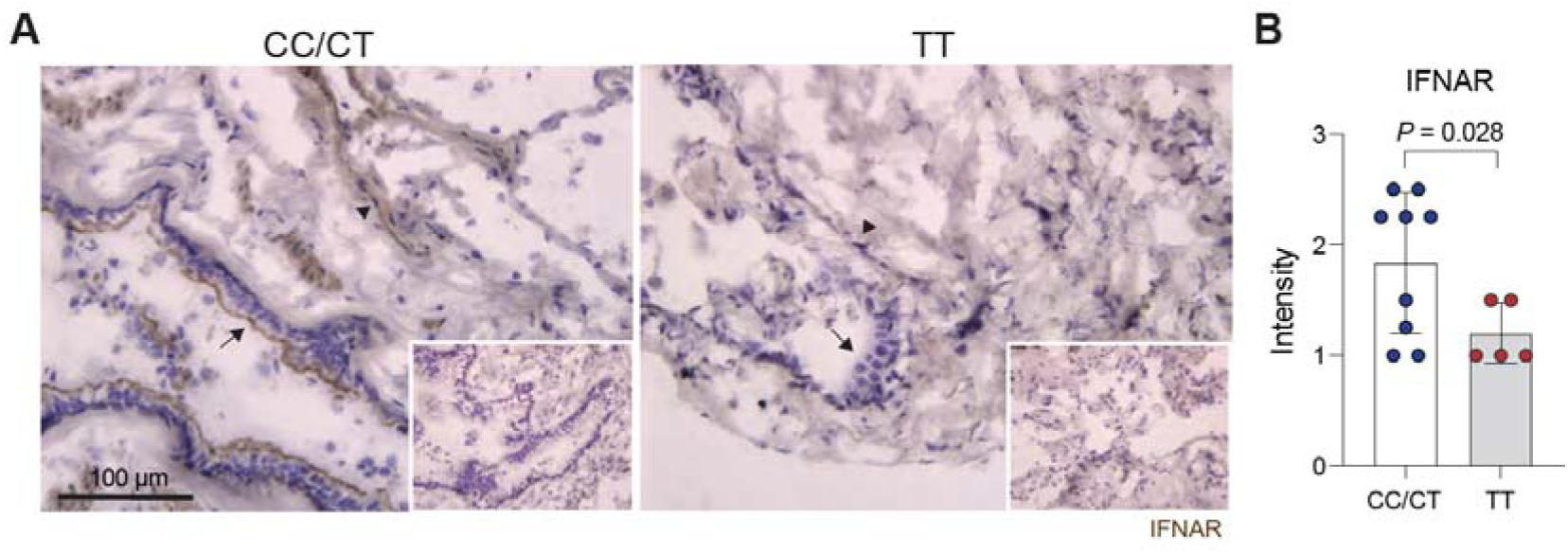
Rs9984273 is associated with the expression of IFNAR. (**A)** Illustrative examples of IFNAR expression in lung specimens from patients with a CT (left panel) and a TT (right panel) genotype. Arrows indicate the alveolar epithelium and arrowheads the vessels. Stained negative controls are shown in the insets. (**B)** Combined expression intensity scores of CC/CT and TT lung specimens (n=9 and 5, respectively). The single CC sample was pooled with CT samples.

**FIG. 3.**
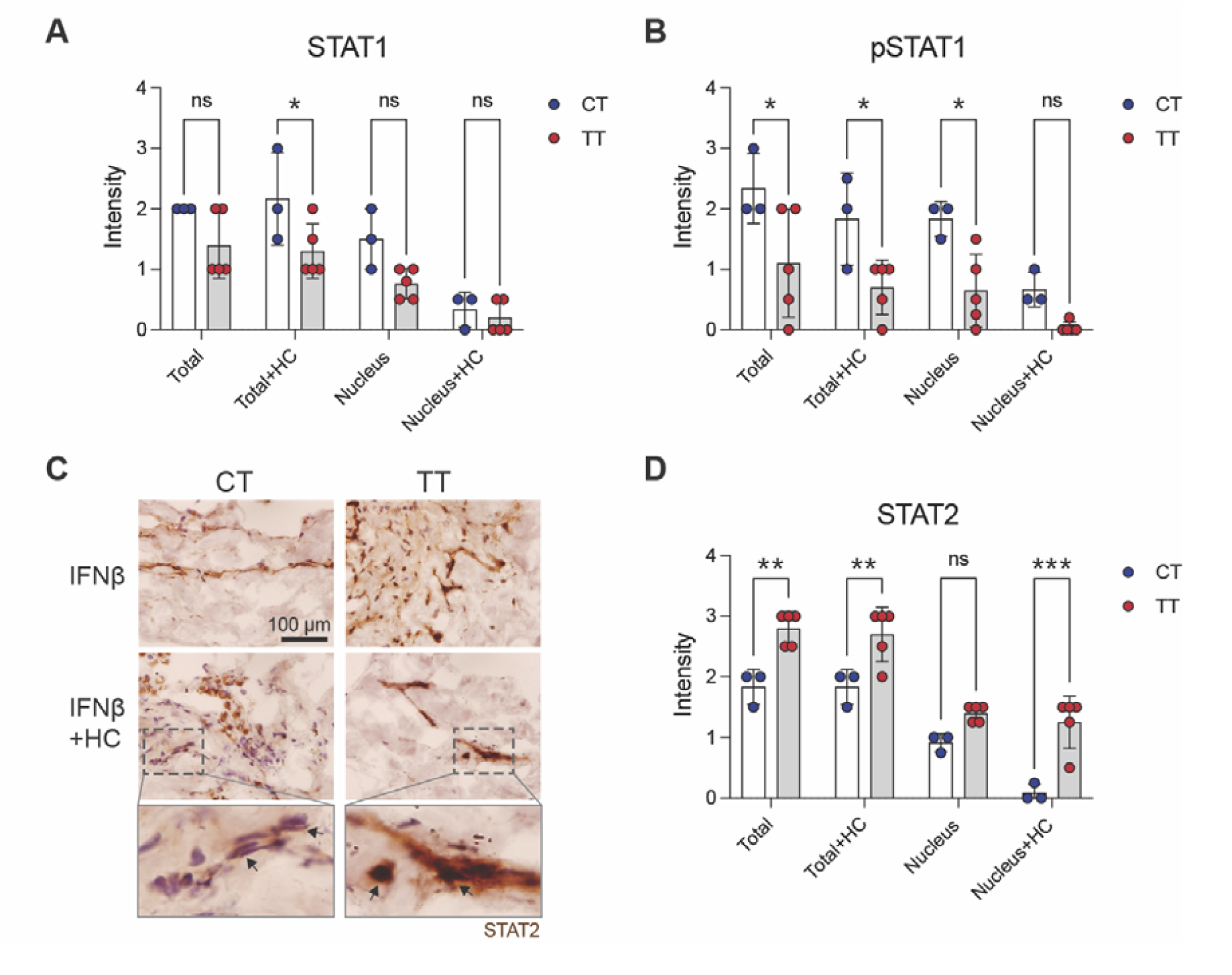
Patients homozygous for the major allele of rs9984273 (TT) have higher STAT2 expression in the lungs than the heterozygous patients with the minor allele (CT) and corticosteroids do not inhibit its nuclear expression in TT patients. **(A**) STAT1 expression in the lung after 4-day culture in the presence of IFN beta with or without hydrocortisone (HC). (**B)** pSTAT1 expression in the same specimens as in A. (**C)** Example photomicrographs showing higher STAT2 expression in a TT patient than in a CT patient and the effect of HC on its nuclear translocation. Most STAT2 remains in the cytoplasm of the CT patients, whereas nuclear expression is prominent in the TT patient. Indicated insets are shown in the bottom row. Arrows. (**D**) Combined results of all patients noting that two CT samples are excluded in the data as the patients were already under glucocorticoid treatment at the time of sample acquisition. Ns, not significant; *P<0.05; **P<0.01; and ***P<0.001

## Rs9984273 is associated with COVID-19 hospitalization

Adhering to our recent clinical and laboratory findings, and the outbreak of the SARS-CoV2, we also explored the role of rs9984273 in COVID-19 through publicly available databases. After all, without a doubt both endogenous and exogenous type I IFNs and steroids play a fundamental role in disease progression and survival concerning COVID-19. Recent genome wide analyses of COVID-19 patients have revealed that SNP rs2236757 in IFNAR2 has a significant impact on disease severity *(24)*, but the functional role of these findings is yet to be explained. Until now, the importance of rs9984273 has not been recognized. Therefore, using the COVID-19 Host Genetics Initiative database, we explored whether there was an association between rs9984273 and disease severity (Table 1). We discovered that the presence of the minor allele of rs9984273 was associated with less hospitalization for COVID-19 (OR 0.96; 95%CI 0.93 – 1.00; P = 0.035), when comparing hospitalized to non-hospitalized COVID-19 patients. Hospitalized patients with COVID-19 also differed from the general public in the context of rs9984273 genotype distribution (OR 0.96; 95%CI 0.93 – 0.98; P = 0.001. Comparing the patient group with the most severe disease (very severe confirmed respiratory COVID-19) to the general public, the rs9984273 polymorphism showed a statistically significant risk association (OR 0.93; 95%CI 0,89 – 0,98; P = 0.006). The number of cohorts involved in the individual comparisons varied from 3 to 60 with no sign of heterogeneity between the cohorts (P > 0.3).

**Table 1.** Associations of rs9984273 to disease severity of COVID-19 in meta-analyses

| Phenotype of cases and controls (database release) | Number of cases with genotype information | Number of controls with genotype information | Odds Ratio (95% confidence interval) | P-value | Number of studies | P-value from Cochran's Q heterogeneity test |
| --- | --- | --- | --- | --- | --- | --- |
| Very severe respiratory confirmed covid vs. not hospitalized covid (release 4) | 269* | 688* | 0.69 (0.46-1.02) | 0.065 | 3 | 0.469 |
| Very severe respiratory confirmed covid vs. general population (release 6) | 6321 | 678513 | 0.93 (0.89-0.98) | 0.006 | 19 | 0.759 |
| Hospitalized covid vs. not hospitalized covid (release 6) | 13582 | 66291 | 0.96 (0.93-1.00) | 0.035 | 20 | 0.313 |
| Hospitalized covid vs. general population (release 6) | 19876 | 1450704 | 0.96 (0.93-0.98) | 0.001 | 33 | 0.291 |
| Covid vs. general population (release 6) | 95644 | 1752242 | 0.99 (0.98-1.00) | 0.048 | 60 | 0.318 |

\*Total number of cases and controls. Subjects involved in the calculation was 679 (distribution of cases and controls used in the calculation was not given in the original data).

SNP rs9984273 is a relatively common polymorphism according to available data. Approximately 45% of people with African origin, 34% of Caucasians and 10% of Asians carry the polymorphism https://www.ncbi.nlm.nih.gov/snp/rs9984273#frequency_tab, which may explain the better resilience of Africa in the current SARS-CoV2 pandemic, as well as vulnerability of the Asian population concerning steroid use for the treatment of COVID-19 *(25)*.. The associations between rs9984273 and mortality in patients with ARDS, as well as hospitalizations with COVID-19 are novel findings as no prior clinical significance has been reported with this polymorphism. We also tested, whether carrying the minor alleles of rs9984273 C and rs2236757 A, correlated with mortality in our ARDS cohort. We found that patients possessing the minor alleles for both genotypes have significantly decreased mortality than those homozygous with the major allele for both genotypes. The lower mortality among the minor allele carriers (n = 51) compared to non-carriers (n = 41) was seen at days 28, 90, 180 and 360, with 28-day mortality of 10% vs. 27% (P = 0.03), 90-day mortality of 16% vs. 37% (P = 0.02), 180-day mortality of 18% vs. 39% (P = 0.02), and 360-day mortality of 20% vs. 39% (P = 0.04), respectively. Moreover, when we tested the presence of these alleles in our lung specimens 80% (4/5) of high IFNAR expressors had both rs2236757 A and rs9984273 C alleles, whereas rs2236757 G was present with rs9984273 C in three out of four low expressors (Fig, 2B). Further, the A allele of rs2236757 alone was not enough for high IFNAR expression as 80% (4/5) of low IFNAR expressors with rs9984273 TT had rs2236757 A. Although rs2236757 in IFNAR2 is associated with severe COVID-19 infection and low IFNAR expression to life-threatening disease *(24)*, rs2236757 and rs9984273 are only in a mild linkage disequilibrium with (r^2^=0.2) in the European population. Nevertheless, they may still significantly co-operate and regulate the expression of IFNAR as four out of five patients possessing both minor alleles had high levels of IFNAR expression.

Both IFN and cortisol responses are both needed in viral induced critical illness, but at different times, IFNs first and steroids later. In most people both endogenous and exogenous steroids block the organ protective effects of IFN beta on the endothelium, if there is no preceding type I IFN response. We have discovered a relatively common genetic polymorphism rs9984273 that prevents GR affecting IFNAR expression, but allows glucocorticoids to shut down STAT1-STAT2 type I IFN signalling. This genetic polymorphism has a significant role in disease states where both type I IFNs and steroids have an impact on mortality. We envision that this genetic polymorphism has evolved and become enriched during previous viral outbreaks and pandemics in humans. Furthermore, his polymorphism may, at least in part, explain the continuous controversy in studies investigating the use of glucocorticoids for ARDS, especially those of viral origin.

## Materials and Methods

### Clinical study populations

The clinical study population has been extensively characterized previously*(18)*. Shortly, The INTEREST trial was a multi-center, randomized, double-blind, parallel-group trial conducted at 74 intensive care units in 8 European countries (from December 2015 to December 2017) that included 296 adults with moderate to severe ARDS. Patients were randomized to receive an intravenous injection of 10 μg IFN beta-1a (144 patients) or placebo (152 patients) for 6 consecutive days. We illustrate the possible interaction between rs9984273 and IFN beta-1a and impact on survival by presenting Kaplan-Meyer survival curves until 1-year follow-up for both study arms with and without concomitant presence of SNP rs9984273. A logistic regression model including age, gender, ARDS severity, Acute Physiology and Chronic Health Evaluation (APACHE) II score, concomitant glucocorticoid use with IFN beta, and SNP status was used to evaluate independent factors related to 28-day mortality.

The association of rs9984273 with survival is reported using the entire study population, but for the IFN gamma analyses presented here only patients with ARDS due to pneumonia or sepsis or pulmonary origin were included as they best resemble COVID-19 ARDS patients. There were 75 of those patients with an available genetic consent and samples available. Daily serum samples were drawn for cytokine measurements. Pro-inflammatory cytokine levels were analysed using Bio-Plex Cytokine Assay by Bio-Rad Laboratories (Hercules, USA). A repeated measurements ANCOVA (RMANCOVA) for change from the baseline in log scale (adjusted for baseline log-transformed value) was used to compare daily IFN gamma and IL-6 levels between the patients according to the SNP rs9984273 status.

### Lung specimens

Lung specimen from 14 different individuals were obtained by post-surgical resection of lung tissue (typically for cancer resections). The lung sections were from lung regions having normal macro- and microscopic appearance and were used with the permission from the ethics authorities at Turku University Hospital (Finland). Several small pieces were cut from all samples and 5-6 pieces/well/condition were cultured for 4 days in 24 well plates containing 1 ml of RPMI medium (supplemented with 10% fetal calf serum, 4 mM L-glutamine, 100 U/ml penicillin, and 100 μg/ml streptomycin), IFN beta (1000 UI/ml FP1201; Faron Pharmaceuticals) or placebo with and without hydrocortisone (40μg/ml, Solucortef, Pfizer). After culturing, all pieces in each well were collected and frozen in OCT. All 14 samples were used for staining IFNAR1 and 5 CT and 5 TT samples for Stat1, pStat1 and Stat2 stainings.

DNA was extracted from 3-5 frozen lung tissue sections (20 μm) with NucleoSpin DNA Rapid Lyse Kit (Macherey-Nagel). TaqMan SNP genotyping assays C_2443264_10 and C_11354003_30 (Thermo Fisher Scientific) for rs9984273 and rs2236757, respectively, were used in genotyping. The reactions were performed as in TaqMan SNP genotyping manuals (ThermoFisher). The plate was run on a QuantStudio 3 Real-Time PCR machine (ThermoFisher) with a run protocol for SNP genotyping as per manual. The results were analyzed using the Applied Biosytems® analysis modules for genotyping in Thermo Fisher Cloud computing platform (ThermoFisher Scientific)

### Immunohistochemistry

Six 5 μm thick frozen sections were cut from lung pieces and fixed with acetone for 5 min. Thereafter, the endogenic peroxidase was blocked using Bloxall Blocking solution (Vector laboratories) for 15 min at room temperature at RT. The first stage antibodies were anti-alpha chain of the IFN alpha/beta receptor (St John’s Laboratory STJ112765) that was used 1:2000 and 1:5000 and anti-Stat1 (1:400, 9175S), anti-pStat1(1:100, 9167S) and anti-Stat2 (1:200, 72604S) all from Cell Signalling. Rabbit Ig (1μg/ml) was used as a negative control. The secondary step was performed with Vectastain Elite ABC rabbit IgG kit (Vector laboratories) and diaminobenzidine was used as a chromogen.

Two different concentrations of the antibody against the alpha chain of the IFN alpha/beta receptor were used to better recognize the differences between the samples. The staining intensity of each sample was blindly scored from 1 to 3. The means of the intensity obtained by different antibody concentration are given. The intensity of the Stat stainings were semiquantitatively scored from 0 to 3 both at the whole cell level and separately in nuclei without knowledge of the patient identity. On average of 20 fields/sample at 200x magnification using a fluorescence microscope (Olympus) were evaluated.

### Description of genetic analyses performed on clinical study subjects

The AmpliSeq® custom panel was used to sequence NT5E (= CD73), IFNAR1, IFNAR2, MCM2, ANGPT2 and RASSF3 of 232 consented study subjects. The panel was designed to cover the whole gene region (exons, introns and untranslated regions (UTRs)) and 3 kb up- and down-stream. Paired-end sequencing with 2 × 300-bp read length was conducted with an Illumina MiSeq® instrument using v3 sequencing chemistry. The quality of the sequencing data was high, and only minor trimming was done to remove low quality bases from the data (<1% of the data). The alignment and variant calling were conducted with the BaseSpace data analysis tool (https://basespace.illumina.com, Illumina). Differences in SNP rs9984273 genotype distribution between deceased and alive ARDS patients were tested using allelic chi-square test using the PLINK software *(26)*. Chi-square test was also used to assess the correlation between the carriers of the minor alleles of rs9984273 and rs2236757 and mortality.

### Binding site analysis

We investigated putative binding sites of GR within the SNP rs9984273 region utilizing the Transfac database (version 2020.3) *(27)* and Encode ChIP-seq data *(22)*. The Match tool in Transfac was used to find matches between the SNP rs9984273 region and the position weight matrices in the Transfac database. The quality of a match between the query region and the matrix was assessed by a matrix similarity score. The HaploReg tool *(28)* was used to find matches between the SNP rs9984273 region and the position weight matrices derived from the Encode ChIP-seq data *(22)*. Only matches that passed the stringent threshold of P < 4^-8^ reported by the tool were considered.

### COVID-19 Host Genetics Initiative database analyses

The COVID-19 Host Genetics Initiative (COVID19-hg) GWAS meta-analyses rounds 4 and 6 summary results were used to identify the association of the SNP rs9984273 with disease severity *(29)*. The meta-analysis across multiple cohorts was performed with fixed effects inverse variance weighting and adjusted for age, sex, and genetic ancestry principal components. Cochran’s Q heterogeneity test was used to examine heterogeneity among the cohorts. SNPs with minor allele frequency less than 0.001 and an imputation quality score (INFO) less than 0.6 were filtered out from each individual study before the meta-analysis. For SNP pruning, the European sub-cohort of the 1000 Genomes phase 3 was used. We inspected the rs9984273 risk association in the available subgroups of very severe respiratory confirmed covid, hospitalized covid, and covid, as compared to not hospitalized covid or general population.

## Data Availability

All data produced in the present work are contained in the manuscript.

## Acknowledgements

The authors want to thank the INTEREST-trial study group for the data analyzed further in this manuscript. We also thank Sari Mäki and Teija Kanasuo for their expert technical help and Dr. Joe Hettinger for valuable comments.

## Author Contributions

Conceptualization: JJ, SJ

Methodology: SK, TH, MH, KE, LE, SJ

Investigation: JJ, SK, SJ

Visualization: MH

Funding acquisition: SJ, LE

Writing - original draft: JJ, SK

Writing – review and editing: all authors

## Data and materials availability

All data are available in the main text.

## References

1. A. Singanayagam et al., Corticosteroid suppression of antiviral immunity increases bacterial loads and mucus production in COPD exacerbations. Nat Commun 9, 2229 (2018).

2. D. B. Coursin, K. E. Wood, Corticosteroid supplementation for adrenal insufficiency. JAMA 287, 236–240 (2002).

3. D. C. Vinh et al., Harnessing Type I IFN Immunity Against SARS-CoV-2 with Early Administration of IFN-β. J Clin Immunol 41, 1425–1442 (2021).

4. J. R. Flammer et al., The type I interferon signaling pathway is a target for glucocorticoid inhibition. Mol Cell Biol 30, 4564–4574 (2010).

5. J. Jalkanen, V. Pettilä, T. Huttunen, M. Hollmén, S. Jalkanen, Glucocorticoids inhibit type I IFN beta signaling and the upregulation of CD73 in human lung. Intensive Care Med, (2020).

6. D. Diez et al., Network analysis identifies a putative role for the PPAR and type 1 interferon pathways in glucocorticoid actions in asthmatics. BMC Med Genomics 5, 27 (2012).

7. S. Y. Ruan et al., Exploring the heterogeneity of effects of corticosteroids on acute respiratory distress syndrome: a systematic review and meta-analysis. Crit Care 18, R63 (2014).

8. T. Thompson, V. M. Ranieri, Steroids are part of rescue therapy in ARDS patients with refractory hypoxemia: no. Intensive Care Med 42, 921–923 (2016).

9. K. P. Steinberg et al., Efficacy and safety of corticosteroids for persistent acute respiratory distress syndrome. N Engl J Med 354, 1671–1684 (2006).

10. J. V. Peter et al., Corticosteroids in the prevention and treatment of acute respiratory distress syndrome (ARDS) in adults: meta-analysis. BMJ 336, 1006–1009 (2008).

11. J. Villar et al., Dexamethasone treatment for the acute respiratory distress syndrome: a multicentre, randomised controlled trial. Lancet Respir Med 8, 267–276 (2020).

12. J. W. Yang, L. Yang, R. G. Luo, J. F. Xu, Corticosteroid administration for viral pneumonia: COVID-19 and beyond. Clin Microbiol Infect 26, 1171–1177 (2020).

13. J. Carlet, D. Payen, S. M. Opal, Steroids for sepsis and ARDS: this eternal controversy remains with COVID-19. Lancet 396, e61–e62 (2020).

14. P. Horby et al., Dexamethasone in Hospitalized Patients with Covid-19. N Engl J Med 384, 693–704 (2021).

15. Y. M. Arabi et al., Interferon Beta-1b and Lopinavir-Ritonavir for Middle East Respiratory Syndrome. N Engl J Med 383, 1645–1656 (2020).

16. I. F. Hung et al., Triple combination of interferon beta-1b, lopinavir-ritonavir, and ribavirin in the treatment of patients admitted to hospital with COVID-19: an open-label, randomised, phase 2 trial. Lancet 395, 1695–1704 (2020).

17. A. J. Spicer, S. Jalkanen, Why Haven’t We Found an Effective Treatment for COVID-19? Front Immunol 12, 644850 (2021).

18. V. M. Ranieri et al., Effect of Intravenous Interferon β-1a on Death and Days Free From Mechanical Ventilation Among Patients With Moderate to Severe Acute Respiratory Distress Syndrome: A Randomized Clinical Trial. JAMA 323, 725–733 (2020).

19. A. C. Kalil et al., Efficacy of interferon beta-1a plus remdesivir compared with remdesivir alone in hospitalised adults with COVID-19: a double-bind, randomised, placebo-controlled, phase 3 trial. Lancet Respir Med 9, 1365–1376 (2021).

20. G. Bellingan et al., The effect of intravenous interferon-beta-1a (FP-1201) on lung CD73 expression and on acute respiratory distress syndrome mortality: an open-label study. Lancet Respir Med 2, 98–107 (2014).

21. G. G. Yegutkin, Adenosine metabolism in the vascular system. Biochem Pharmacol 187, 114373 (2021).

22. P. Kheradpour, M. Kellis, Systematic discovery and characterization of regulatory motifs in ENCODE TF binding experiments. Nucleic Acids Res 42, 2976–2987 (2014).

23. J. Nan, Y. Wang, J. Yang, G. R. Stark, IRF9 and unphosphorylated STAT2 cooperate with NF-κB to drive IL6 expression. Proc Natl Acad Sci U S A 115, 3906–3911 (2018).

24. E. Pairo-Castineira et al., Genetic mechanisms of critical illness in COVID-19. Nature 591, 92–98 (2021).

25. J. Liu et al., Corticosteroid treatment in severe COVID-19 patients with acute respiratory distress syndrome. J Clin Invest 130, 6417–6428 (2020).

26. S. Purcell et al., PLINK: a tool set for whole-genome association and population-based linkage analyses. Am J Hum Genet 81, 559–575 (2007).

27. V. Matys et al., TRANSFAC and its module TRANSCompel: transcriptional gene regulation in eukaryotes. Nucleic Acids Res 34, D108–110 (2006).

28. L. D. Ward, M. Kellis, HaploReg: a resource for exploring chromatin states, conservation, and regulatory motif alterations within sets of genetically linked variants. Nucleic Acids Res 40, D930–934 (2012).

29. C.-H. G. Initiative, The COVID-19 Host Genetics Initiative, a global initiative to elucidate the role of host genetic factors in susceptibility and severity of the SARS-CoV-2 virus pandemic. Eur J Hum Genet 28, 715–718 (2020).

